# Anti-CD20 antibodies in relapsing multiple sclerosis: Protocol for a systematic review and network meta-analysis

**DOI:** 10.1101/2024.01.12.24301256

**Authors:** Cristian Eduardo Navarro, Simón Cárdenas-Robledo, Jaiver Macea, Natalia A Ortíz-Cano, Ivan D Florez

**Affiliations:** School of Medicine, Universidad de Antioquia. Medellín, Colombia; Grupo de Investigación E.S.E Hospital Emiro Quintero Cañizares. Ocaña, Colombia; Centro de Esclerosis Múltiple, Hospital Universitario Nacional (CEMHUN). Bogotá, Colombia; Departamento de Medicina Interna, Universidad Nacional de Colombia. Bogotá, Colombia; Research Group Experimental Neurology, Department of Neurosciences, KU Leuven. Leuven, Belgium; Pharmacy Department, Hospital Pablo Tobón. Medellín, Colombia; Department of Pediatrics, University of Antioquia. Medellín, Colombia; School of Rehabilitation Science, McMaster University. Hamilton, Canada; Pediatric Intensive Care Unit, Clínica Las Americas-AUNA. Medellín, Colombia

## Abstract

**Background:** Multiple sclerosis is a chronic disease of the central nervous system characterized by autoimmune demyelination. Various immunomodulatory medications are available for its treatment, among which anti-CD20 monoclonal antibodies have demonstrated superior efficacy compared to other disease-modifying therapies. However, there is a lack of direct comparison between the four available anti-CD20 monoclonal antibodies. Therefore, this study aims to systematically assess the relative efficacy and safety of anti-CD20 monoclonal antibodies for treating relapsing multiple sclerosis.

**Materials and Methods:** We will conduct a systematic review of phase IIb and phase III clinical trials of anti-CD20 monoclonal antibodies for the treatment of relapsing multiple sclerosis, following the Preferred Reporting Items for Systematic Reviews and Meta-Analysis guidelines. A comprehensive review of the Cochrane Central Register of Controlled Trials; MEDLINE; Embase; ClinicalTrials; International Clinical Trials Registry Platform; OpenGray; and MedRxiv will be performed. Two independent reviewers will select titles, abstracts, and eligible full texts to execute the data extraction. The risk of bias will be assessed with the Cochrane RoB 2 tool. Data on pre-specified outcomes will be analyzed for the selected articles using a random-effects network meta-analysis with a frequentist framework. Summary statistics, along with 95% confidence intervals will be presented, and the effectiveness and safety of each intervention will be ranked using the surface under the cumulative ranking curve. A missing data and a sensitivity analysis will be conducted. We will use the GRADE approach to assess the certainty of the direct, indirect, and network estimate for all outcomes.

**Results and Conclusion:** The results will provide evidence of the relative efficacy and safety of anti-CD20 monoclonal antibodies for the treatment of relapsing multiple sclerosis.

**Systematic review registration:** PROSPERO registration number CRD42023437996

## Introduction

Multiple sclerosis (MS) is a chronic inflammatory disease of the central nervous system (CNS) that affects millions of people worldwide [1,2]. MS is characterized by the loss of myelin, which leads to impaired nerve function and disability. Demyelination was traditionally considered to be caused by autoreactive T lymphocytes [3], but the role of B lymphocytes in the MS pathogenesis has been increasingly recognized during the last two decades [4]. As a result, B cell depleting anti-CD20 monoclonal antibodies have emerged as promising disease-modifying therapies (DMTs) for MS [5–8]. Early phase 1 and phase 2 trials showed reduction in magnetic resonance imaging (MRI) activity (in the form of new/enlarging FLAIR/T2 lesions and gadolinium-enhancing [Gd+] lesions) and a discrete reduction in the long-term clinical outcome in people with inflammatory activity in MRI after treatment with rituximab [9,10].

Three new anti-CD20 monoclonal antibodies have been approved by regulatory agencies for the use in relapsing-remitting MS (ocrelizumab, ofatumumab and ublituximab), and primary progressive MS (ocrelizumab). All three have been studied in phase 3 randomized controlled trials (RCTs) [11–14]. These have had active comparator and placebo arms and have shown to be effective in reducing the disease relapsing rates. but, to our knowledge, no direct comparison between each other, or with rituximab has been published. As a result, to date it is not clear whether the anti-CD20 are similar in their beneficial effects and potential harms. Since head-to-head RCTs comparing the interventions among them may take years if not decades, the alternative is to conduct evidence synthesis methods that allow us to estimate the relative efficacy using indirect evidence in the cases when direct evidence is not available.

## Materials and Methods

### Study registration

This protocol is reported in accordance to the Preferred Reporting Items for Meta-Analyses Protocols (PRISMA-P) guidelines (S1 Table) [15] and has been registered in the International Prospective Register of Systematic Reviews (PROSPERO) (registration number CRD42023437996).

### Objective

Assess the comparative efficacy, safety, and tolerability of anti-CD20 monoclonal antibodies for treating relapsing MS, through a systematic review and network meta-analysis (NMA).

## Eligibility criteria

### Type of studies and participants

We will include phase 2b and phase 3 placebo-controlled, and active-comparator RCTs. Active comparators will include any other anti-CD20 monoclonal antibody or any other DMT, such as, but not restricted to: alemtuzumab, cladribine, dimethyl fumarate, fingolimod, glatiramer acetate, interferon beta-1a, interferon beta-1b, pegylated interferon beta-1a, natalizumab, ozanimod, siponimod, and teriflunomide. Patients 18 years of age and older with MS confirmed with 2010 or 2017 McDonald criteria [16,17], and with a relapsing-remitting course according to the current classification will be included [18]. Studies published in English or Spanish will be included, with no restriction regarding the year of publication. Quasi-randomized, open-label, cluster and cross-over trials will not be included.

### Type of interventions

The interventions to be assessed are anti-CD20 monoclonal antibodies (ocrelizumab, ofatumumab, rituximab, ublituximab) irrespective of dose, dosage form, route of administration, administration frequency, duration of treatment, and previous exposure to any active interventions.

### Type of outcomes

Considering the prioritized outcomes suggested by the MS Core Outcome Set [19], the primary efficacy outcome will be the annualized relapse rate (ARR) defined as the number of confirmed relapses of multiple sclerosis per participant-year. The secondary efficacy outcomes will be the proportion of patients with disability progression confirmed at 12 week– and 24 week-(confirmed disability progression defined as an increase from the baseline EDSS [Expanded Disability Status Scale] score of at least 1.0 point or 0.5 points if the baseline EDSS score was >5.5) [20], and the number of Gd+ lesions and the new/enlarging FLAIR/T2 lesions.

The primary safety outcome will be the proportion of patients with any adverse event (AE) as defined by the study authors. The proportion of patients with serious adverse events (SAE) and infusion-related reactions will be considered as secondary safety outcomes.

## Information sources and search strategy

### Data sources and search strategy

We will search the following databases from their inception: Cochrane Central Register of Controlled Trials, MEDLINE, Embase, ClinicalTrials (https://clinicaltrials.gov), International Clinical Trials Registry Platform (https://www.who.int/clinical-trials-registry-platform/the-ictrp-search-portal), OpenGray (https://easy.dans.knaw.nl/ui/home), and MedRxiv (https://www.medrxiv.org). We will also review the reference lists of included studies and relevant reviews for additional studies not included after the main searches.

To build the search strategy a list of MeSH terms and keywords related to “anti-CD20”, “Antibodies, Monoclonal”, “drug-modifying therapy”, “multiple sclerosis”, “Multiple Sclerosis, Relapsing-Remitting”, “ocrelizumab”, “ofatumumab”, “rituximab” and “ublituximab” will be considered. We will use the validated RCT filter created by the McMaster University Health Information Research Unit for MEDLINE and Embase through the Ovid platform. These filter provide a good balance between sensitivity and specificity [21].

The preliminary and the final search strategy will be based on the recommendations provided by the Cochrane Handbook for Systematic Reviews of Interventions [22], tailored to be specific for each database. A draft of the MEDLINE search strategy is shown in Table 1. We will export all the searches results to Mendeley® version 1.19.8 and will remove duplicates.

**Table 1.**
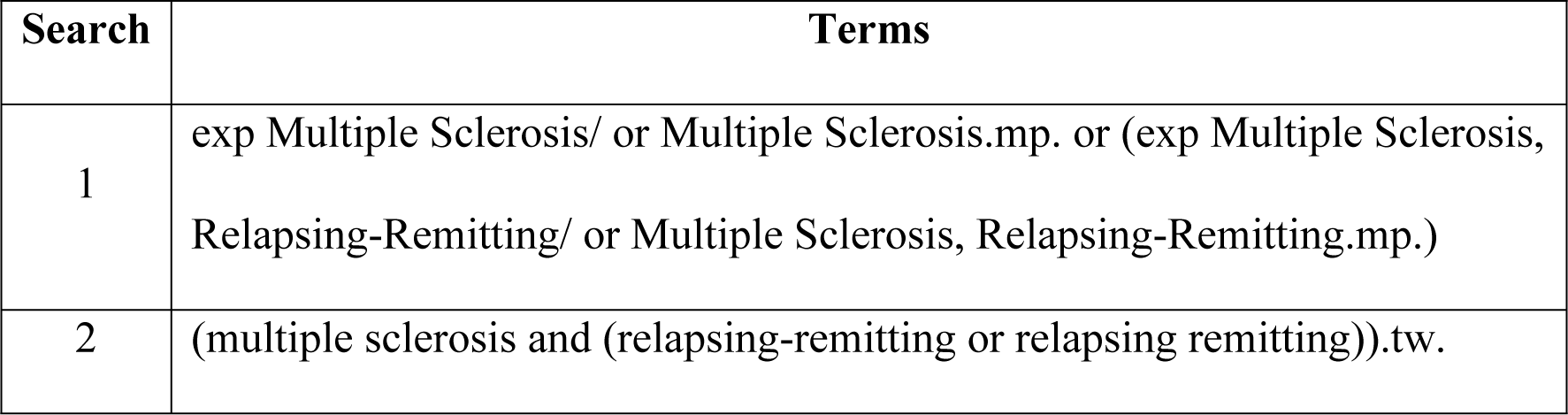
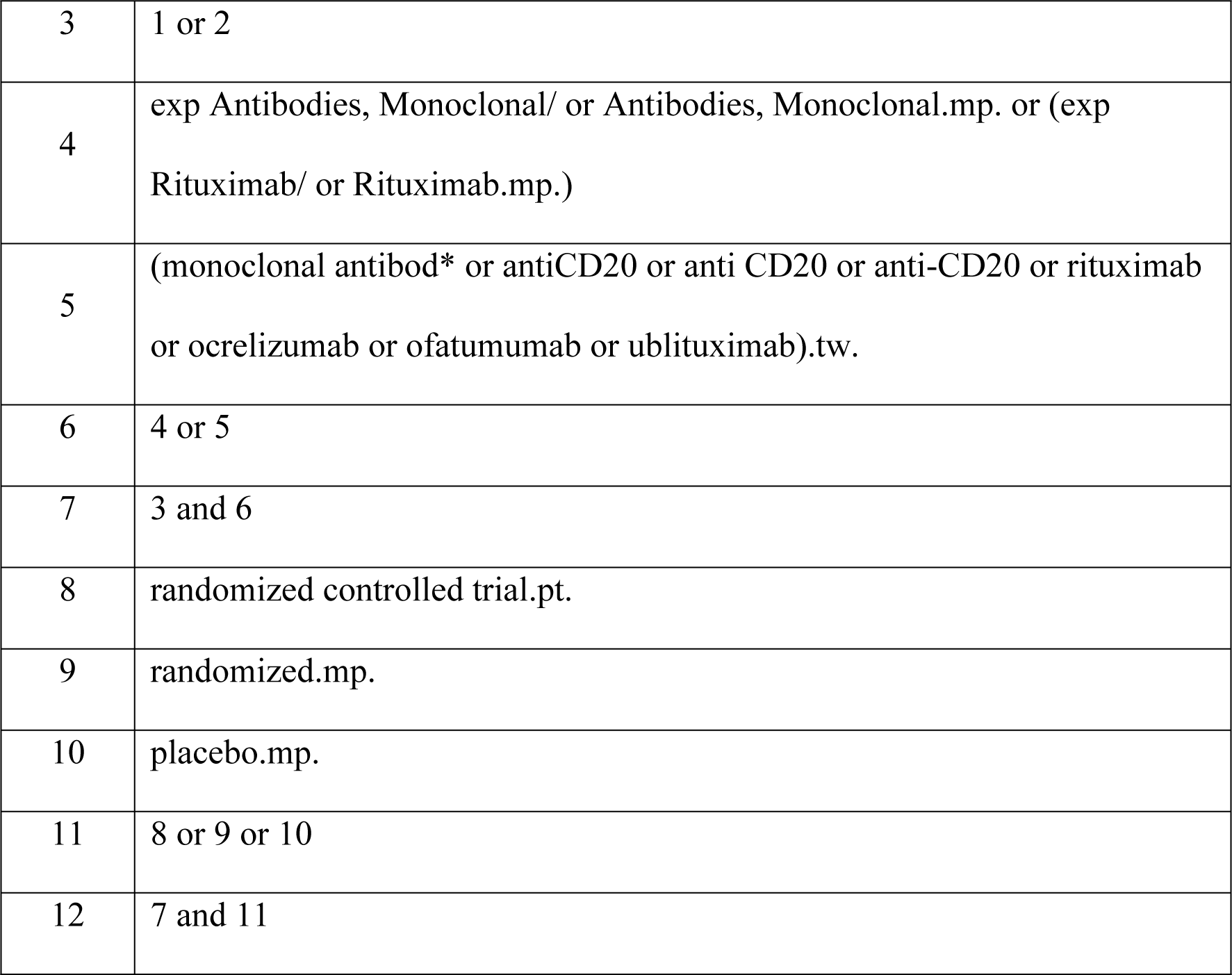
Draft of search strategy in MEDLINE (via Ovid)

## Selection process, data collection and management

### Study selection

Two independent reviewers (CEN, JM) will independently and in duplicate screen all he retrieved titles and abstracts to determine their eligibility according to a prespecified and piloted screening format. We will retrieve the full-text references of studies that at least one review author considers to be eligible during the screening. The full-text records will then be reviewed to determine the eligibility criteria using a prespecified and piloted screening format. Disagreements between the reviewers will be resolved by consensus. In the event of persistence of discrepancy between the two reviewers a third evaluator will determine the eligibility (IDF). We will report a PRISMA flow diagram of included and excluded articles [23]. The *“Characteristics of excluded studies”* section will detail the primary reason for exclusions.

### Data extraction

Two authors (CEN, JM) will extract data from the eligible studies using a standardized data extraction form independently and in duplicate. This will include 1) characteristics of the study: trial registry ID number, first author, year of publication, single or multicenter study, study design, inclusion/exclusion criteria, sample size per arm, recruitment and sampling procedures used, enrolment start and end dates, length of participant follow-up, methods used to prevent and control for confounding, source(s) of funding, authors’ financial relationship and other potential conflicts of interest; 2) characteristics of participants: age in years, sex, time since symptom onset, disease duration, disability measured by EDSS, pre-intervention mean number of relapses, mean baseline number of FLAIR/T2 and Gd+ T1 lesions, previous DMT used; 3) details of the intervention and comparator: intervention, dose, formulation, frequency and mode of application, duration of administration; and 4) outcome results: outcome, timing of outcome measurements, number of participants randomly assigned and included in the analysis, number of participants who withdrew, were lost to follow-up or were excluded, number of any adverse event occurred, number of serious adverse events and number of infusion-related reactions. For dichotomous outcomes we will extract the number of events and number of participants randomized and for continuous outcomes, the mean and standard deviations (SD) per arm.

### Risk of bias assessment

Two independent reviewers (CEN, JM) will assess the risk of bias in duplicate of the included studies using the Cochrane RoB 2 tool [24], which includes the following domains: bias arising from the randomization process, bias due to deviations from intended interventions, bias due to missing outcome data, bias in the measurement of the outcome; and bias in the selection of the reported result. For each domain, we will reach a risk of bias judgment, assigning one of three levels to each domain: low risk of bias; some concerns; or high risk of bias. The overall risk of bias domain will be classified as: ‘low risk of bias’ when all domains are judged as low risk, ‘some concerns’ when at least one domain is some concern but was not high risk for any domain and ‘high risk’ if at least one domain was high risk or if multiple domains were judged as some concerns [24]. Any discrepancies will be resolved through discussion or by a third reviewer (IDF). Studies will not be excluded based on the risk of bias assessments, but we will conduct sensitivity analyses to explore the potential effects of high risk of bias in the meta-analyses. Industry funding will be considered as a potential source of bias [25].

### Data synthesis

We will summarize trial and population characteristics, using descriptive statistics. For each separate outcome, we will present a network plot of available trial data to be included in the analysis. Network nodes will represent interventions compared within trials and edges will represent trials directly comparing the corresponding interventions. We are primarily interested in the following nodes: rituximab, ocrelizumab, ofatumumab and ublituximab. We anticipate some degree of variability in the way in which the interventions are administered. Namely, some interventions may have been evaluated at different doses and regimens of administration. Therefore, if we identify heterogeneity at the pairwise comparisons levels and this seems to be related to differences in the doses or methods of administration, we will split some interventions into two or more nodes to represent these differences. The size of each node will be proportional to the number of participants in the underlying intervention, and each line edges will be weighted according to the number of studies comparing the interventions it connects. To identify the most influential comparisons in the network, we will present the contribution matrix describing the contribution of each direct estimate to the entire network of trials [26].

### Statistical analysis

We will conduct a network meta-analysis (NMA) using a frequentist framework. We will use a random-effects model to estimate the treatment effect for each drug compared to placebo and with other DMT [27,28]. We will also compare the efficacy and safety of each drug to each other using a network meta-analysis (NMA) using multivariate distributions to allow for the simultaneous analysis of multiple interventions and account for correlations induced by multi-arm trials [29]. We will calculate odds ratio (OR) for dichotomous outcomes and mean differences (MD) or standardized mean differences (SMD) for continuous outcomes, both with their corresponding 95% confidence intervals (95%CIs). If a study does not provide enough data for conducting the statistical analysis, we will try contacting the study authors. If this is not feasible, or we do not obtain any response from them, and the required data is crucial for the analysis (eg, mean results for continuous outcomes) we will exclude the study from the pooled data and present the information narratively. In case authors provide variability measures in a different format (eg, standard error, or 95%CI), we will follow the recommendations by the Cochrane Handbook of Systematic Reviews to estimate the required standard deviation [22]. When studies report median and ranges or interquartile ranges, we will use the methods by Wan to estimate the best mean and SD [30].

### Pairwise meta-analysis

We will conduct pairwise meta-analysis of the available direct comparisons. We will conduct an inverse variance random-effects meta-analysis to estimate overall effect using OR for dichotomous data and MD or SMD for continuous outcomes. We will assess heterogeneity by visually comparing clinical and methodological characteristics of the included studies and by visual inspection of study effect sizes and their variation in the forest plots. We will also calculate the Chi square test for heterogeneity, and the I^2^ statistic. We will consider an I^2^ value >50% to be indicative of substantial heterogeneity [31]. We will perform subgroup or sensitivity analyses to explore the potential reasons behind substantial or considerable heterogeneity, namely, the potential effect modifiers.

*A priori*, we have identified the following variables might be effect modifiers: age, relapses in the 12 months prior to randomization, disease severity according to EDSS, presence of GD+ lesions in MRI, and previous use of other DMT. We hypothesize that patients with younger age, high number of relapses at baseline, milder disability, the presence of Gd+ lesions and no prior treatment show larger effect. To assess the credibility of an apparent effect modification, we will use the Instrument for assessing the Credibility of Effect Modification Analyses (ICEMAN) [32].

### Network meta-analysis

We will conduct an inverse variance random-effects meta-analysis to estimate overall effect sizes, that is respectively ORs or MDs/SMDs for dichotomous or continuous outcomes, along with their corresponding 95%CIs, under the assumption that different trials are estimating different but related true effects. We will use the Hartung-Knapp-Sidik-Jonkman method to calculate a 95%CI for the overall effect size to handle meta-analyses with a small number of studies [33]. We will rank the drugs according to their efficacy and safety based on P-scores.

For a connected network of trials, we will conduct a random-effects NMA [34], if we consider that the assumptions of transitivity and consistency are justifiable. For transitivity, we are assuming that participants are equally likely to be randomized to any of the included interventions and that trials are sufficiently similar across comparisons with respect to their effect modifiers distribution. We will consider the same effect modifiers described above as potential causes of heterogeneity. To explore variability in intervention definitions, we will split them into separate nodes according to the doses administered in the included trials.

The statistical manifestation of intransitivity can create inconsistency between direct and indirect evidence. We will assess consistency within each network using both global and local statistical approaches. We will evaluate each network using the design-by-treatment interaction test, and accounting for multiple sources of inconsistency due to disagreements in trials with different designs [35–37]. We will also evaluate consistency in each loop of interventions in each network using the loop-specific approach, and between each direct and indirect evidence using the node-splitting approach [38,39]. We will plot all consistency estimates and their 95%CIs to make inferences about the presence of inconsistency locally in the network.

### Missing data and sensitivity analysis

We will conduct sensitivity analyses to assess the robustness of our findings for the primary outcomes. First, we will conduct an analysis by excluding studies with an overall high risk of bias. If the results are very different from the analyses including all studies, we will prioritize the results obtained from the studies with ‘low risk’ and ‘some concerns’ analyses for our primary findings. Second, we will perform an analysis excluding trials for which we imputed data. Lastly, we will perform an analysis using the intention to treat denominators, in the case of trials that had data imputed. In case of missing participant data, we will capture the type of data imputation used by the trial authors in our extraction form, to assess the appropriateness of the data imputation method. For missing outcome data, we will try to contact authors for more information. If the trial authors do not respond, we will consider alternatives such as imputation approaches. If the missing data from the studies are significant, we will consider this in the ‘Bias due to missing outcome data’ criterion of the RoB 2 assessment.

We will use comparison-adjusted funnel plots to assess the likelihood of reporting bias and small-study effects for the NMA when at least 10 studies per outcome are available [40–42]. In the case of funnel plot asymmetry, we will explore possible sources of it, including poor methodological quality in smaller trials, true heterogeneity, non-reporting biases, and baseline risk differences.

In the cases in which the key assumptions are not met (transitivity and consistency), we will provide only pairwise estimates and a narrative description of the results. We will use R software version 4.3.2 (The R Foundation, Wien, Austria) for the analyses.

### Summary of findings and assessment of the certainty of the evidence

We will use the GRADE approach to assess the certainty of the direct, indirect, and network estimate for all outcomes. Two review authors (CEN, JM) will independently assess the certainty of the evidence for each outcome. Where necessary, we will resolve any disagreements between authors through discussion or by consultation with a third review author (IDF).

For the direct evidence, the assessment starts by considering evidence from RCTs to be of high certainty, and the body of summarized evidence is assessed against five criteria: study limitations (risk of bias) [43], inconsistency [44], indirectness [45] imprecision [46], and publication bias [47]. For the “study limitations” criterion, we will use the overall risk of bias assessment judgment obtained from the RoB 2 tool [24]. For the specific NMA GRADE assessment we will use the specific approach that considers intransitivity [48] and incoherence criteria [49] and we will apply the approach to draw conclusions from the NMA sing a minimally-contextualized framework [50].

### Ethical consideration

This study does not require ethical approval as it is a secondary analysis of published data. We will disseminate the results of this study through publication in a peer-reviewed journal and presentation at relevant conferences.

## Discussion and conclusion

This protocol outlines a network meta-analysis comparing the efficacy and safety of anti-CD20 monoclonal antibodies for treating relapsing multiple sclerosis.

Anti-CD20 monoclonal antibodies have shown remarkable results in terms of efficacy for treating MS, which has led to their inclusion in an application to the World Health Organization essential medicines list [51]. However, their relative efficacy between them has not been assessed in controlled trials to date. Several RCTs aiming to demonstrate the non-inferiority of rituximab in comparison to ocrelizumab are underway [52–55], but results are expected for 2025 at the earliest. Further, to the best of our knowledge there are no studies underway comparing the efficacy of the newer anti-CD20 monoclonal antibodies. Therefore, the results of this study will provide valuable information to guide clinical decision-making, to inform future research in this area, and to develop new health economic evaluations related to these DMTs [56].

We expect difficulties for the completion of this study, for we anticipate that we will find a limited number of RCTs, with high heterogeneity which probably will affect the results of the meta-analysis.

## Data Availability

Deidentified research data will be made publicly available when the study is completed and published

## Supporting information

**S1 Table. PRISMA-P (Preferred Reporting Items for Systematic review and Meta-Analysis Protocols) 2015 checklist**

